# Robot dance: a city-wise automatic control of Covid-19 mitigation levels

**DOI:** 10.1101/2020.05.11.20098541

**Authors:** Paulo J. S. Silva, Tiago Pereira, Luis Gustavo Nonato

**Affiliations:** University of Campinas; University of Sao Paulo

## Abstract

We develop an automatic control system to help to design efficient mitigation measures for the Covid-19 epidemic in cities. Taking into account parameters associated to the population of each city and the mobility among them, the optimal control framework suggests the level and duration of protective measures that must be implemented to ensure that the number of infected individuals is within a range that avoids the collapse of the health care system. Compared against other mitigation measures that are implemented simultaneously and in equal strength across cities our method has three major particularities when:

**Accounts for city commute and health infrastructure:** It takes into account the daily commute among cities to estimate the dynamics of infected people while keeping the number of infected people within a desired level at each city avoiding the collapse of its health care system. **City-specific control:** It allows for orchestrating the control measures among cities so as to prevent all cities to face the same level control. The model tends to induce alternation between periods of stricter controls and periods of a more normal life in each city and among the cities. **Flexible scenarios:** It is flexible enough to allow for simulating the impact of particular actions. For example, one can simulate the how the control all cities change when the number of care beds increases in specific places.

Therefore, our method creates an automatic dance adjusting mitigation levels within cities and alternating among cities as suggested in [9]. This automatic dance may help the city economy and orchestration of resources.

We provide case studies using the major cities of the state of São Paulo given by using estimates on the daily mobility among the cities their health care system capacity. We use official data in our case studies. However, sub-notification of infected people in Brazil is notoriously high. Hence the case study should not be considered as a real world policy suggestion. It high sub-notification is taken into account, the optimal control algorithm will suggest stricter mitigation measures, as also shown in the case studies. Surprisingly, the total duration of the protocol for the state is barely affected by the sub-notification, but the severity of such protocols is strengthened. This stresses a twofold implication, first, the protocol depends on high-quality data and, second, such optimal and orchestrated protocol is robust and can be adjusted to the demand.

## 1 Introduction

After the first cases of Covid-19 appeared in Brazil on February 25, the state of São Paulo was the first to adopt social distancing on March 26, after the first school closures on March 17. The social distancing protocol played a major role to control the rapidly increasing number of cases and thereby protecting the health care system [11]. While an early implementation of an aggressive protocol of social distancing is effective, it has three major drawbacks:

**Long implementation:** A social distancing protocol, or any other mitigation strategy, must be implemented over many months. Otherwise, it only causes a delay in the spreading of the virus while the peak of infected individuals remains virtually unchanged. This means that *short* social distancing only postpones the main issues.

**Simultaneous implementation:** Since the whole state follows the same protocol, cities in the countryside must start it early, while the disease is still in the first stages and they still can cope with the number of cases, ignoring that the peak of cases does not happen at the same time for all cities.

**Homogeneous protocol across cities:** The protocol ignores the distinct roles cities play economically as well as their structure such as hospital and critical care beds.

Now, deep into a seven weeks long protocol, the control measures will be slowly relaxed starting on May 31st. This means that in the State of São Paulo, and in fact in the whole country, another social distancing protocol must be implemented again in the near future. Research shows that intermittent social distancing protocol will need to be implemented in the next two years [6] most likely in an intermittent way, dancing between periods of mitigation and normal life [9]. This is troublesome as the economies of developing countries cannot cope with such extended lockdowns [10].

There is an urgent need to develop tools that may help policy-makers to device intelligent lockdown systems that predict where, how long, and to what extent the social distancing protocol must be implemented. Probably such decisions must be taken at a city level, as each city will have its own economical, geographical, and infrastructural needs.

Our project addresses this challenge by developing a tool that suggests the level of mitigation needed for different cities at different moments, thereby aiming to alleviate the disruption to the activities while protecting the city’s health system. However, we must emphasize that such approach depends on the data quality to estimate the current status of the pandemic. In this sense, it is also a proof of concept of what can be achieved with high quality widespread testing and good estimates of population movement to better understand the disease dynamics. The three major benefits of the algorithm are:

1. It takes into account the city health system capacity to predict the optimal measures.
2. It is flexible to incorporate control policies such as preventing full country lockdown.
3. It provides a city specific timing and extent of control implementation.

## 2 Model and optimization

### 2.1 An SEIR model that with workforce travels

Individuals can be in one of the following states Susceptible (S), Exposed (E), Infected (I), and Recovered (R) [14]. We take into account the fact that during the control protocol most travels between cities are due to the workforce: the workers leave their city to work in another one and return during the evening. Thus, we divide the susceptible population of the city into the population that stays within it and the population that commutes to work.

*During the day:* During working hours the available population of the *i*th city can be increased by the inflow of people workers (resp. outflow) giving an effective population which can be larger (resp. smaller) than the city’s own population. We assume for now that susceptible individuals of the *i*th city going to the *j*th can only be infected there during the day.

*During the night:* As workers come back to their city the model reduces to the standard SEIR model where cities do not interact and have their own population.

*Full day cycle:* Because workers move in and out at well defined times we approximate the movement dynamics as a step function. So all workers move out and come back at the same time. Let *α* be a square pulse modeling this cycle, so that *α*(*t*) = 1 when *t* is the range of working hours (here from 8am to 6pm) and zero otherwise. A representation of the daily cycle is exhibited in Figure 1.

**Figure 1:**
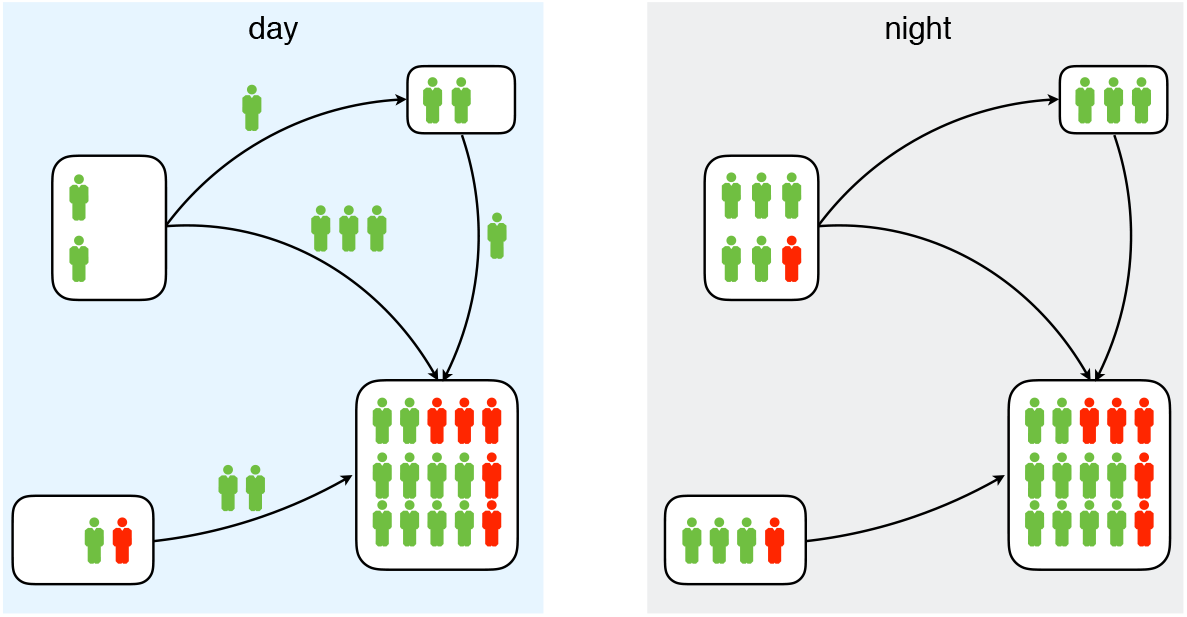
Daily dynamics of workers traveling to nearby cities. The boxes represent the city’s total population. Workers travel to adjacent cities and change the effective population of the city during the day. These workers can get infected at their workplace and serve as a source of infection during the night. For better visualization, we show only susceptible (green) and infected (red).

*Population splitting from data:* The model needs a commuting matrix *p_ij_*(*t*) that contains daily accumulated percentage of inhabitants of *i* that travels to *j*. Here, *p_ij_* may be updated daily. We are assuming that there is no correlation between the states and travels. See further details in App. A.

### 2.2 The control model

In the SEIR models above, the parameter *r_t_*, the reproduction number at time *t* is usually a constant *r*_0_ that represents how many people a sick person infects if all other individuals are susceptible. The main objective of control measures is to decrease *r_t_* to a lower level, bringing the spread to a desired level. Therefore the problem of choosing such *r_t_* can be seen as an optimal control problem where the *r_t_* is the control variable that drives the disease trajectory to an acceptable, or even a desirable, state.

This control problem can then be approximated by an optimization model after we devise a discretization scheme for the ODE. Since the SEIR model is not stiff, it is possible to use simple explicit integration methods and still get good approximations of the trajectory. We used the modified Euler’s algorithm, also known as Heun’s method [4], that computes explicit approximations of the trajectory *S_t_, E_t_, I_t_,R_t_* for each day given the initial values *S*_1_, *E*_1_,*R*_1_, *I*_1_ and the reproduction values *r*_t_. Since the integration method is simple, it can be described as a constraint set of a regular optimization problem. This problem can then be enriched with extra constraints and an objective function arriving at a conceptual model that must be solved by an optimization algorithm. Since the SEIR model is nonlinear, the final optimization problem will also be nonlinear. Finally, in order to take into account the coupling between the dynamics of the epidemic among all cities, described in 2.1, a single problem has to solve the differential equations and choose the target reproduction level of all cities simultaneously.

In our first implementation, we are adding a constraint to the variable *I_t_* for each city at each day. By doing this we try to control for the maximum level of infection that is allowed to avoid saturation of the health system. Another group of constraints tries to avoid abrupt changes of the control *r*_t_, either fixing it in a time window or adding a total variation regularization [2].

As objective we aim to balance between to main goals: trying to enforce the most relaxed control measures, by minimizing the mean deviation of *r_t_* and *r*_0_, and alternating stricter controls in different cities. Such objectives must be carefully balanced to take into account the economical importance of each city for example. At this first implementation we are using the square of the population as our main measure for the importance of each city. In a real world application such weights must be given by the decision-makers taking into account different economical and even political aspects.

Finally the optimization problem was coded using the modeling language JuMP [3]. This is a Julia [1] package that allow us to easily describe the model and that efficiently interfaces with different solvers. It also takes care of efficiently computing the first and second derivatives needed by the solvers. Such flexibility is essential, as the model will evolve with time and should be easy to adapt. To actually solve the optimization problem we are using Ipopt [13], a high performance large scale nonlinear optimization solver, with the MA97 linear equation solver [5] that is able to exploit multiple cores.

For more details and specific formulas for the constraints and objective see Appendix B.

## 3 Case Studies – Optimizing the isolation protocol

We discuss case studies involving possible scenarios of Covid-19 control in the major cities of the state of São Paulo. Our control depends on the data quality, ranging from the city health infrastructure to the evolution of the number of cases. The latter is notoriously compromised in Brazil [12]. Thus, for sake of comparison, we will show three scenarios: the current social distancing protocol, optimal control taking into account the office data, and finally, optimal control taking into account sub notification. In all cases we use mobility matrices estimated from anonymized mobile phone data [8].

### Current social distancing only

This scenario uses *official government data*, which is sub-notified [12]. Assuming that social distancing policy started in March 24 is kept until the end of June with the current effectiveness, we find that the peak of infected people in São Paulo is reached in early July, while for the other cities in the metropolitan region of São Paulo will face the peak of the epidemic outbreak in August, as shown in Figure 2. São Paulo city would have more than 2% of its population infected for about two full months, from early June to early August. An *optimistic* estimate (including campaign hospitals) shows that the São Paulo health system has a maximum capacity of 1.5% of the population infected.

**Figure 2:**
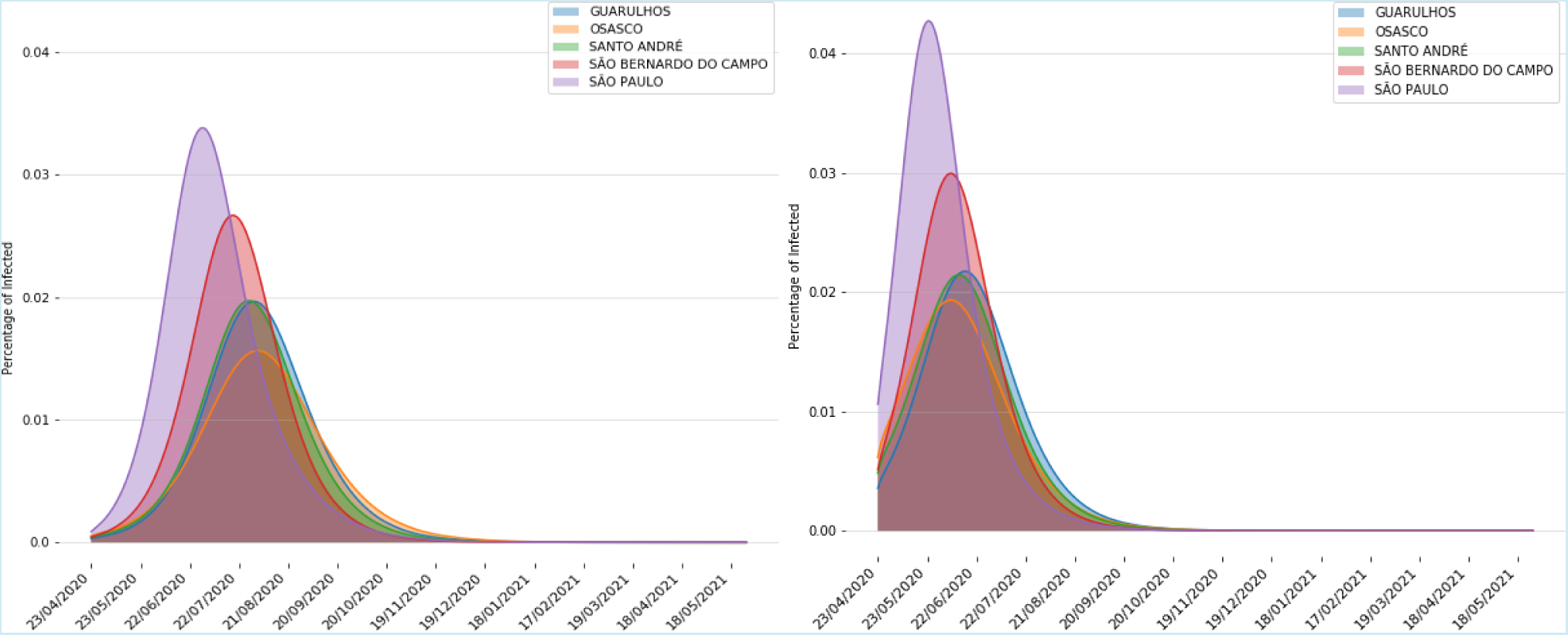
Prediction of Covid-19 spread under the current social distancing level. We take into account commuting among cities and consider the *official data*. The prediction assumes that the social distancing protocol implemented in the state of São Paulo would continue until July and with the same effectiveness. The infected population in the city of São Paulo would be above 1.5% for over two months causing a full collapse of the health system.

### Optimal and flexible control

As we discussed in the introduction our approach has three major advantages that we illustrate bellow by adjusting them to following measures

1. We fixed the maximal percentage of infected to be 1.5% for the city of São Paulo and 0.7% for the countryside. This corresponds to an optimistic estimate of the health care capacity.
2. We implement a control protocol where cities aim for distinct control measures to prevent a full state lockdown and thus keep the local state economy running.
3. For the official data, our protocol is allowed to seek for controls with a minimal reproduction number 1. When taking into account sub-notification we implement an initial control phase where the minimal reproduction number is 0.89. This is necessary because some cities already start above the threshold level for the infected, described above, while others we reach it in a few days due to the initial dynamics.

As a consequence, each city follow the protocol according to their own need and will have their starting and stopping time of control which takes into account city to city commutes.

*Office government data:* In the left inset of Figure 3, we show the results of the above implementation using *official data* released by the Brazilian government. We provide control measures in six levels according to how strict they need to be ranging from no control (white) to severe (red). Each city will have its control target to protect its health system. The optimal control provides distinct protocols according to the city role in the State. For instance, Guarulhos has severe control which is gradually decreased until the city is back to open, whereas a nearby city Santo André swings between a period of severe control to open. The optimal control solutions have three remarkable aspects:

- The control releases São Paulo earlier than other cities. As São Paulo health system can serve twice the number of infected people than other cities, the control to end much earlier there.
- Countryside cities such as Araçatuba, Ribeirão Preto e Rio Preto, could start the control two weeks later than the cities in the Greater São Paulo. They follow the control until late September when they alternate between two weeks of lock-down and no control.
- Most cities in the Metropolitan area of São Paulo can start the intermittent fortnightly distancing periods much earlier, by mid-August. During the period of intermittent control, the cities, mainly in the countryside, do not stop at the same time, an import aspect to keep the economy always running in part of the state during the epidemic outbreak.

**Figure 3:**
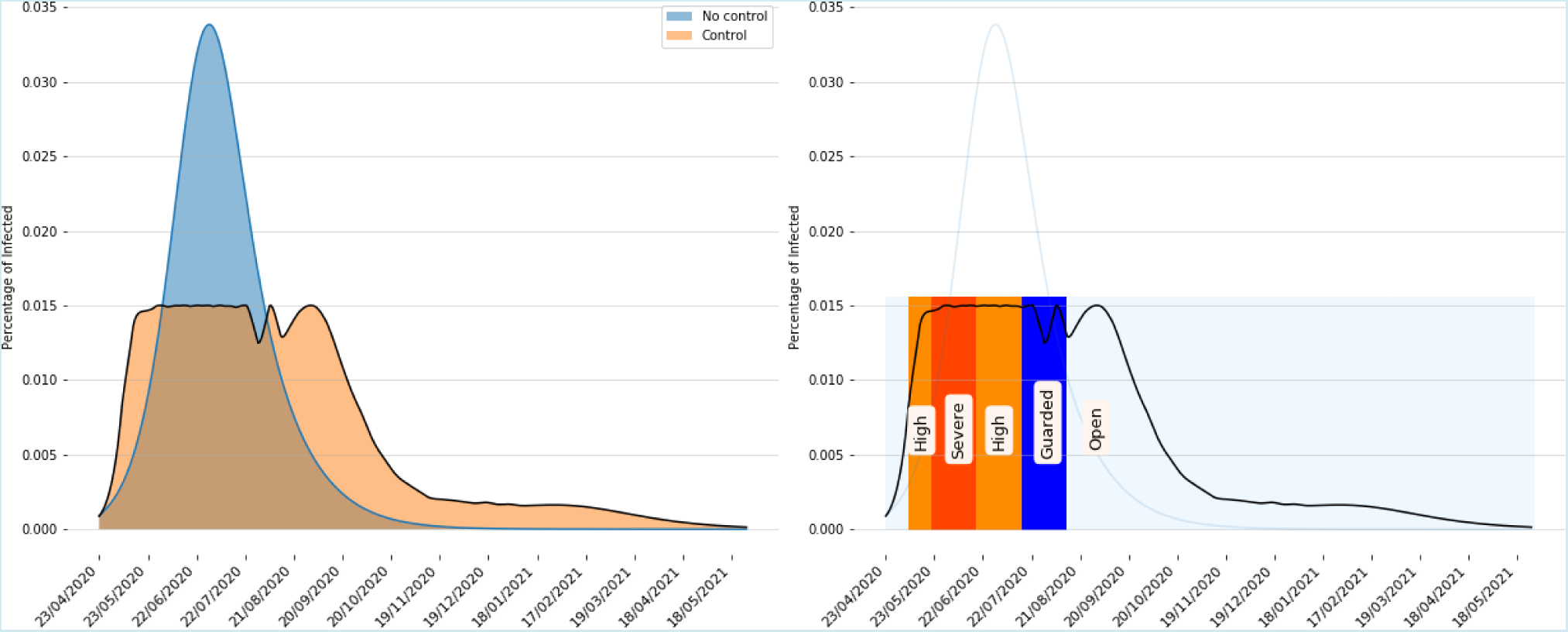
Impact of the optimal control protocol implemented in the State of São Paulo. The control protocol does not allow for the infected population in the city of São Paulo to surpass 1.5% and in the countryside 0.7% (this would roughly correspond to take campaign hospitals into consideration). In the left inset, we show the situation with social distancing only (blue) and with optimal control (beige) where the infected population never exceeds 1.5% as set in the control. In the right inset, our approach provides levels of control for the city of São Paulo. This prediction performed using the official government data.

*Taking into account sub-notification:* In the right inset of Figure 3, we run the optimal control taking into account the strong sub-notification in the official data [12]. Clearly, there is no margin to manoeuvre and the severe control starts straight away. However, increasing the number of infected twelve-fold only changes slightly the duration of the optimal control protocol.

Notice that by taking into account sub-notification severe control is applied more often. In particular, severe control must be imposed during June, after that the level of control is progressively relaxed, starting the opening period from mid-August. Notice that if a proper level of control is imposed in specific time intervals the duration of control increases in only about two weeks, but at a much more moderate level in such extra weeks.

## Discussion

We start the discussion emphasizing the importance of reliable data to convert the scenarios simulated by our approach in concrete actions. Using official data, the left inset of Figure 4 shows that after one month of social distancing in São Paulo state, the control could be avoided in May for most cities in the countryside of São Paulo state if the only objective is to preclude the collapse of the health care system. If the simulation takes into account sub-notification, such relief is not feasible. In fact, cities in the countryside of the state as well as in the metropolitan area of São Paulo must undergo a *severe* mitigation during the first two weeks of May. Cities in the countryside of the state can only relax the control in the second fortnight of May. Therefore, without reliable data it is complicated to simulate scenarios with low degree of uncertainty, making evident the need for testing the population in a broad scale in order to take secure actions.

**Figure 4:**
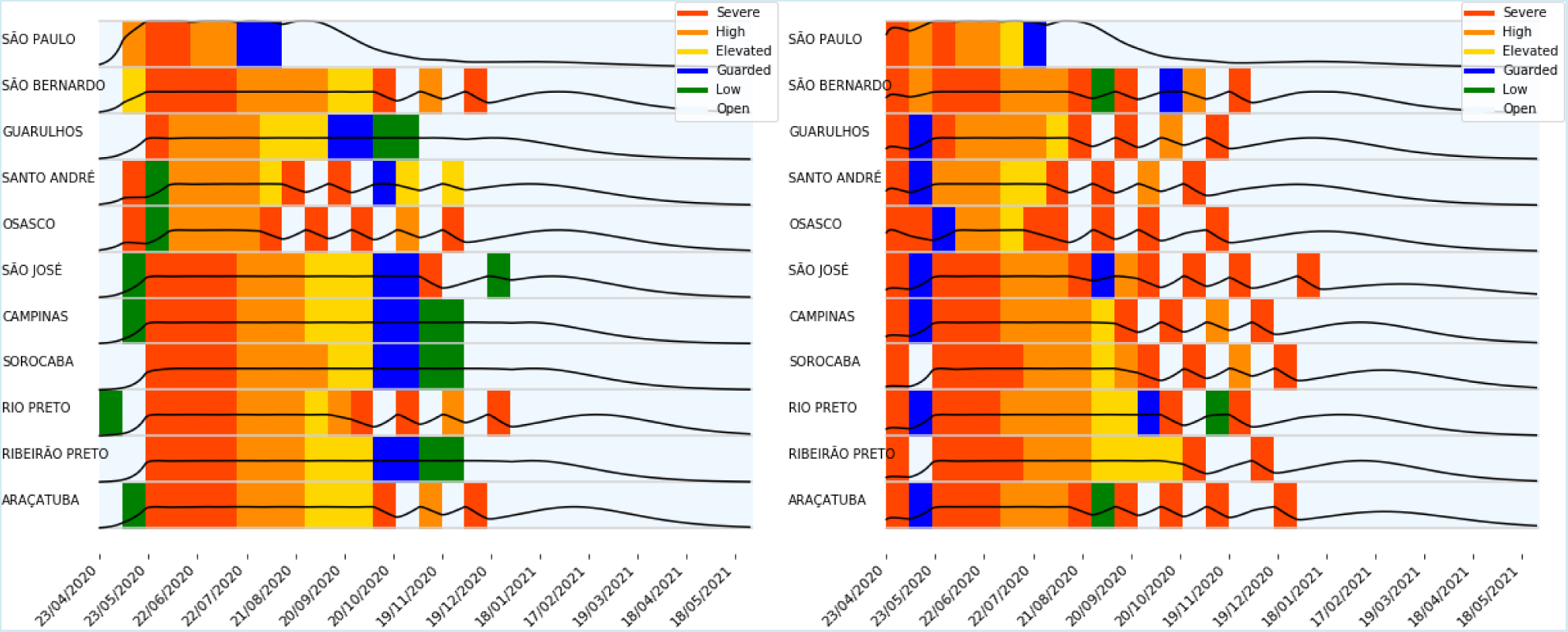
Prediction of optimal control for major cities in the state of São Paulo. The protocol changes every two weeks and optimizes the level control and the timing to apply the control measures. Thus each city can meet its own needs to curb the spreading. In the left inset, we show the optimal control taking into account the official data. In the right inset, we take into account a sub-notification.

Figure 5 presents another scenario simulated by our methodology. In this example we take into account that the epidemic outbreak ends first in São Paulo than in other cities. Therefore, from September on, the simulation makes 50% of the hospital beds from São Paulo available to the other cities. The larger number of beds available for other cities allows an increase in the number of infected people in the other cities, shortening the period of rigid control, mainly for cities in interior of the state, in about 45 days, from early October to mid August. Notice how the black curves goes above halfway after September, showing a larger number of people is infected in each city, but still under the admissible limit. The intermittent period of severe control are also shortened in about one month.

**Figure 5:**
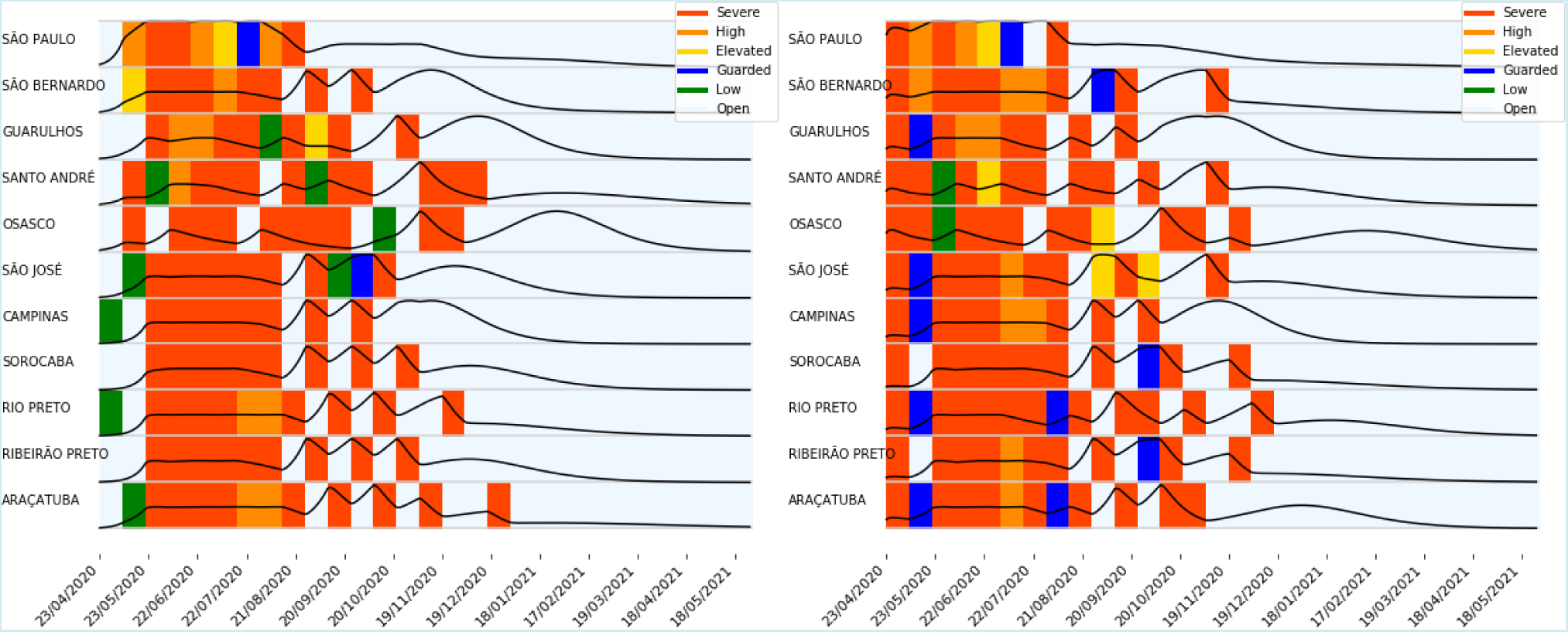
Using the health care infrastructure of the city of São Paulo to assist others. The protocol changes every two weeks and optimizes the level control and the timing to apply the control measures. In late August, when São Paulo has already passed the peak of epidemic outbreak, half of its hospital beds are made available to assist other cities, in the interior and in metropolitan area, shortening the time interval of severe control policy in those cities. Left: Official data; Right: Assuming sub-notified data.

The case studies discussed above shows the power of the proposed methodology in simulating different scenarios and henceforth enabling decision-making agents to analyze multiple alternatives so as to balance city-wise health system capacity and economic factors over São Paulo state.

### Level of control and actions adapted to a city level

The outcome of the optimization is the reproduction number the city must attain during the implementation phase of the protocol.

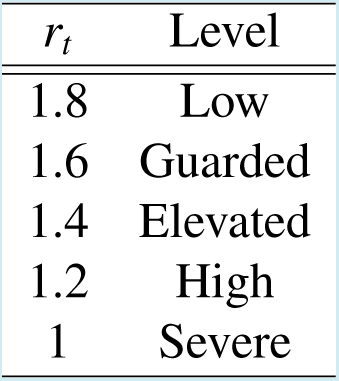

| $r_t$ | Level |
| --- | --- |
| 1.8 | Low |
| 1.6 | Guarded |
| 1.4 | Elevated |
| 1.2 | High |
| 1 | Severe |

It is mandatory that we translate these variables into actions that policy-makers can implement at a city level. The reproduction number is sensitive to the density of the population and services level.

## 4 Conclusions

We developed a city based flexible control protocol for Covid-19 mitigation and used cities in the state of São Paulo as a case study. The protocol takes into account infrastructure, care beds and mobility data from the cities to plan an optimal control strategy. Here, we focused on strategies that avoid total State lockdown and thus keep the economy running and open opportunity to use medical resources in an efficient way.

The recommended mitigation taking into account the official government data is is clearly different to the recommendations when we take sub-notification into account. Surprisingly, the total duration of the control remains almost unaffected. As we showed, the quality of the recommendations depends strongly on the data. Our control can be updated dynamically and can integrate new data on the fly.

## Data Availability

They may be obtained upon request

## Acknowledgements

We are partially supported by the Serrapilheira Institute, Royal Society London, CEPID-CeMEAI (FAPESP 2013/07375-0), FAPESP (2018/24293-0), and CNPq (304301/2019-1, 303552/2017-4, 302836/2018-7). We thank Luisa Souza Moura, David Cairuz da Silva, Marina Fontes Alcantara Machado, and Joao Pedro Rodrigues Mattos for gathering and preprocessing the data used in our simulation.

## A Model derivation

We consider a set of *K* cities with fixed population

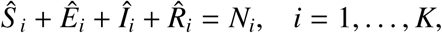

where *N_i_* is the total population. We divide the susceptible population of city *i* into classes as

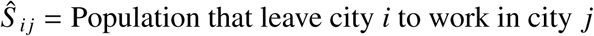

and similarly for the other states *E,I*, and *R*.

**During the day:** The effective population

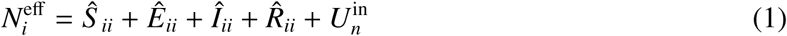

is assumed constant throughout the day but can present daily variations. Here 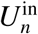 is neighbor inflow

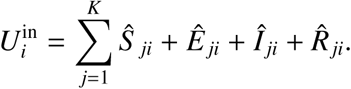

Susceptibles of the *i*th city going to the *j*th can only be infected there. Thus,

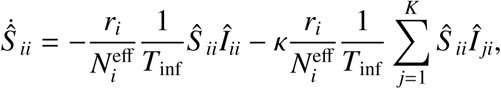

where *κ* is the fraction of infected that can still commute and hence spread the disease elsewhere. For now, we consider *κ* = 0 and then

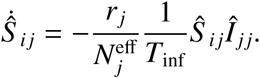

As we consider the absolute population the reproduction number is scaled according to the available population, which is assumed constant during the day. Next we do two steps:

i. Normalize the population *S. + E_i_ + I. + R_i_ =* 1, and thus,

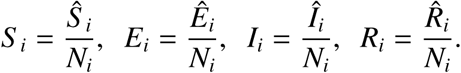
ii. Write in an equation for 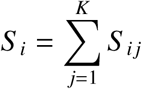 yielding

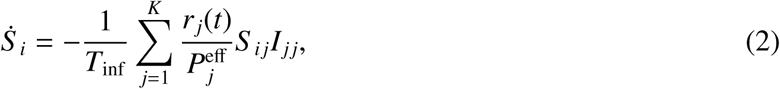

where

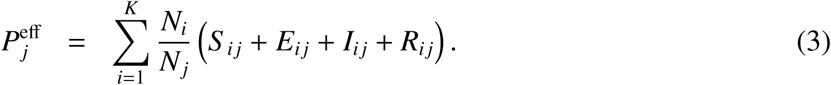

**Population splitting from data:** The fraction of the population *S _ij_* can be estimated using a matrix that describes the mobility between the cities made available in [8]. It has at each entry an estimate of

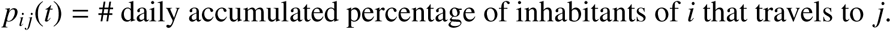

Assuming that there is no correlation between the states and travels, we can assume that the population traveling population is uniform across states that is

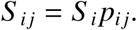

**During the night**. As workers come back to their city the model reduces to the standard SEIR model where cities do not interact and have their own population.

**Full day cycle**. Because the work dynamics is mainly stereotyped and workers move in and out at well defined times we will approximate the dynamics movement as a step function. So all workers move out and come back at the same time. Let *α* be a square pulse modeling this cycle, so that *α*(*t*) *=* 1 when *t* is the range of working hours (here from 8 am to 6 pm) and zero otherwise. Thus the model can be written as

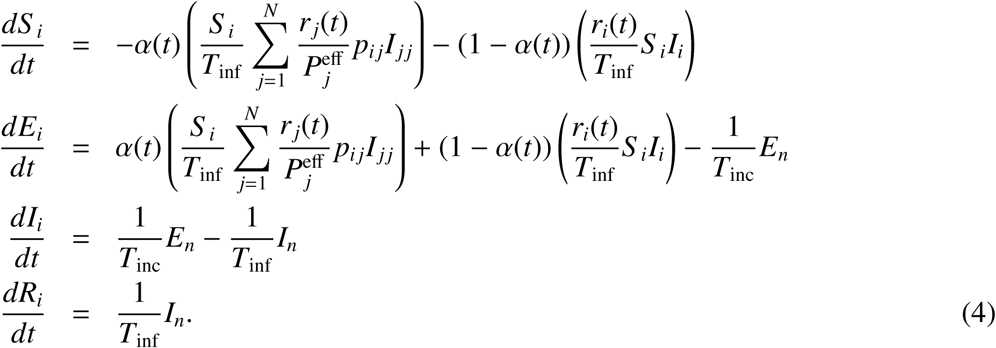

Here we use T_inc_ = 5.2 and T_inf_ = 2.9, following [14].

## B Optimization model

As described in Section 2.2, we approximate the solution of the control problem by solving an optimization model. This model has the decision variables *S_i,t_, E_i,t_*, *R_i,t_*, *I_i,t_, r_i,t_* for *i =* 1,…, *K* and t = 1, …, *T*. The *S, E, I, R* variables represent the normalized SEIR states for all cities *i* at each day *t*. As for *r_i,t_*, it represents the respective target reproduction number that acts as the control variable. The value *K* is the number of cities and *T* is the desired time horizon for the simulation.

We then solve

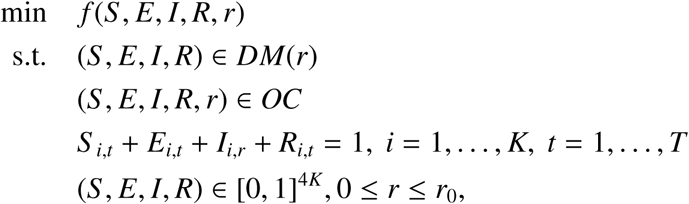

where *DM*(*r*) is a discretized version of the the SEIR model for a given control *r*, as described in the last section, *OC* represents an abstract version of the operational constraints, that we detail below, and *r*_0_ is the natural reproduction number of Covid-19, that we approximated by 2.5 from [7].

We used the modified Euler method, also known as Heun’s method [4], to define *DM*(*r*). This is an explicit second order Runge-Kutta scheme that showed precise enough for our simulations. We used daily time steps.

The operational constraints can be divided in two groups. First we add constraints ensuring that the controls have to remain constant in a time window. In the case studies the time window was 14 days. This is desirable, as it is impossible to rapidly change the behavior of large populations. Next we have constraints that limit the *I* variable at each day, using an estimate of number of intensive care units available for each city. Another possibility would be to add this constraint aggregating the capacity by regions or even for the whole State, simulating situations where there is a single queue for the intensive care units.

Finally, the objective function *f* is composed of two terms that try to balance between two conflicting goals. The first term minimizes

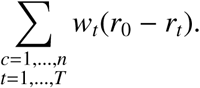

The goal here is to try to avoid the use of mitigation measures, allowing normal life and economic activity. The weight *w_t_* balances the importance associated with keeping normal life at each city. In our case studies we used the square root of the city population to define these weight as a way to give more importance to large cities, if possible.

The second term try to avoid employing the same controls for two different the cities at the same time

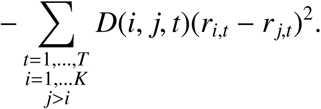

The weights *D*(*i, j, t*) try to measure the importance keeping the cities *i* and *j* under different controls at each time *t*. We used a value proportional to the square root of the population of the smaller city and made it vanish by the end of the simulation, when the epidemic should be in natural decline. In both cases we used values proportional to the square of the population in avoid the large city of São Paulo to fully dominate the decisions.

Finally one has to find weights that balance between those two (conflicting) constraints in order do define the single objective *f*.

This framework is actually quite flexible. It allows for different terms to be added or deleted from the operational constraints *OC* or to the objective *f*. The only constraints that are essential are the ones in *DM*(*r*), since they approximate the SEIR dynamics.

